# Association between altitude and death from COVID-19 in subjects with diabetes mellitus during the first wave: cross sectional study of the National Death Index

**DOI:** 10.1101/2024.02.12.24302726

**Authors:** Raúl Manuel Ambrosio-Ojose, Cesar Josué Casquero-Zambrano, Tery Vasquez-Hassinger, Marlon Yovera-Aldana

**Affiliations:** Carrera de Medicina Humana, Facultad de Ciencias de la Salud, Universidad Científica del Sur, Humana, Lima, Perú; Grupo de Investigación en Neurociencias, Efectividad Clínica y Salud Pública, Universidad Científica del Sur, Lima, Perú

**Keywords:** (MESH): COVID-19, Mortality, diabetes mellitus, Altitude, Peru, risk

## Abstract

**Background:** In high Andean areas, there is greater insulin sensitivity which may be a protective factor against complications in subjects with diabetes mellitus.

**Objective:** Determine the association between altitude of residence and death from COVID-19 in deaths with diabetes mellitus in Peru during the first wave..

**Methods:** We carried out a cross-sectional analysis of deaths registered in the National Death System of Peru (SINADEF in Spanish). We selected Peruvians with diabetes mellitus identified by presenting the diagnosis in any of the six boxes on the certificate. The dependent variable was death from COVID-19 as the basic cause of death, located in last place among causes A, B, C and D according to the Pan American Health Organization. The independent variable was the altitude of residence, categorized as less than 1 500 m a.s.l, 1 500 to 2 499 m a.s.l. and greater than 2 500 m a.sl. Through a multilevel analysis by geographic region and using a Poisson regression, we obtained the risk ratios of death from COVID-19 according to the altitude of residence. We adjusted by individual and contextual variables.

**Results:** We included 16 406 deaths with diabetes mellitus between March-December 2020. 34.3% died from Covid19 and 9.7% came from areas above 2 500 m. The proportion of deaths from COVID-19 of those with residence altitude above 2 500 m was 20% lower compared to residents below 1 500 m (RR: 0.80; 95% CI: 0.70 – 0.91; p<0.001), adjusted for individual and socioeconomic factors. Its influence is also shown as the altitude changes every 100, 250, 500 and 1000 m a.s.l., through multilevel analysis.

**Conclusion:** A higher altitude of residence is associated with a lower proportion of deaths from COVID-19 in people with diabetes mellitus during the first wave in Peru. The study contribute to expanding knowledge of the effects of altitude with respect to mortality in people with diabetes mellitus in a context of a highly contagious and virulent infectious disease.

## Introduction

Peru presented the highest mortality rate from COVID-19 worldwide during the first wave in 2020. Reaching a rate of more than 70 per 100 000[1] and an excess of deaths of 103% compared to 2019 [2]. Fragmented health system with limited budget and inefficient primary care, despite prolonged confinement, led to these serious outcomes[3] .

One of the factors associated with mortality from SARS-Cov-2 is type 2 diabetes mellitus (DM). About 45% of patients with SARS-CoV-2 presented DM at the beginning of the first wave worldwide.[4] It also increases mortality by 0.8 times compared to people without diabetes [5]. An overexpression of angiotensin converting enzyme (ACE) receptor has been discovered on beta cells in the pancreas of subjects with DM This would cause a greater entry of the virus, cell damage and a potential crisis of hyperglycemia that initiates the cascade of acute complications. [6].

In the middle of the first wave, it was postulated that SARS-CoV-2 disease was less prevalent at higher altitudes [7]. In others Latin American countries, altitude also was associated with decrease in morbidity and mortality from COVID-19 [8] A lower expression of ACE is due to chronic hypoxia which causes a lower viral load and a decrease in systemic inflammation [9]. Thus, the impact of altitude on the cause of death is controversial [10,11]. However, in high Andean areas, there is greater insulin sensitivity and a lower prevalence of diabetes mellitus, which may be a protective factor against complications. [12]

Considering that Peru had one of the highest percentages of people living above 2 500 m and one of the highest incidences of diabetes in the world [13] .We aimed to evaluate the association between altitude of residence and death from COVID-19 in deceased Peruvians with DM using data from the National System of Deaths from March to December 2020.

## Materials and methods

### Study design

We conducted a cross-sectional study of data from the National System of Deaths of Peru (SINADEF, initials in Spanish) of the Government of Peru. SINADEF registers the characteristics and cause of death of all Peruvians in national territory and abroad since 2015. Online registration is carried out by any Peruvian doctor and is essential for death certification. The analysis period ranged from March 1 to December 31, 2020.

### Population

We included subjects with diabetes mellitus, without specifying the type of diabetes. It could be like cause of death or comorbidity. ICD-10 code for diabetes mellitus must be in any of the six available boxes on the death certificate. Codes used are in the Supplementary Table 1. Likewise, we excluded Peruvians who died abroad, incomplete data or unclear cause of death.

### Variables

#### Death due to COVID 19

We defined cause of death as COVID 19, if it was present as the underlying cause on the death certificate. [14] Later called the SINADEF criterion and did not require confirmatory tests or symptoms. [15] The Peruvian death certificate, according to guideline of the 10/100 list of the Pan American Health Organization, establishes three causes of death: direct, intermediate and basic. The basic cause of death is defined as that which initiates the chain of pathological events that led directly to death. Which is located in last place among causes A, B, C and D. [16] The ICD-10 codes used were (U07.1) for confirmed disease and (U07.2) for probable disease. Likewise, B34X and B97X codes and records without ICD-10 but written diagnosis such as “SARS CoV-2”, “COVID-19” or “coronavirus” were classified as “other codes [17].

The underlying cause was reclassified as infectious, noncommunicable, external injury, and undetermined. The ICD-10 codes that make up each of these categories can be seen in the *Supplementary Table 2*.

#### Altitude of residence and other variables

Altitude was classified as less than 1500 m, between 1500 and 2499 m, and greater than or equal to 2500 m. [18] The natural region of each district was also described according to Javier Pulgar Vidal’s classification as follows: Lima and Callao, rest of the coast, Sierra and Selva [19]. We described the Peruvian monetary poverty of 2018 in quintiles according to the National Institute of Statistics and Informatics (INEI in Spanish). The original report presented the poverty percentage of the 1874 districts of Peru in descending order. We reclassified by quintiles from 1 to 5. Quintile 1 was the one with the highest percentage of poverty. INEI defines the monetary poverty as those whose per capita spending is below the value of a basket of products (food and non-food such as housing, clothing, education, health, transportation, etc.). that allows the minimum needs to be satisfied. It includes monetary and non-monetary expenses such as self-consumption, self-supply, payments in kind, transfers from other households and public donations. This data was prepared using the theorem of Elbers et al and was calculated by combining the results of a household survey with representative regions and census information. [20]. A summarized version of the poverty values for each quintile can be seen in the supplementary table 3.

From data of the death certificates, we described sex (female, male), age group (0 to 11.9; 12 to 18; 18 to 29.9; 30 to 59.9 and ≥60 years old), education level (illiterate, primary, secondary, higher, not registered), health services insurance (General Population, Employee, Armed or Police Forces, Private and uninsured) and place of death (health center, home, public road / in transit / workplace, unknown). Finally, we describe the presence of arterial hypertension (yes/no), obesity (yes/no), cancer (yes/no), coronary heart disease (yes/no), and cerebral stroke (yes/no). The definitions used were based on ICD-10 coding or written diagnoses.

#### Procedures

The database was downloaded on January 12, 2021 from the National Data Open Platform of the Government of Peru [21].

The diagnosis of COVID-19 or comorbidities was based on the finding of the respective ICD-10 codes. For those who did not have ICD-10 coding, manual searches for words or groups of letters that approximated the diagnosis were carried out using the filters of the Microsoft Excel spreadsheet. The ICD-10 codes and texts used are detailed in the **Supplementary Table 1**.

The diagnosis of the underlying cause of death was made by a physician with experience in clinical epidemiology and management of the death database. Taking as priority the diagnosis of last place according to the PAHO provision for the completion of death certificates and the evaluator’s criteria.

The SINADEF database does not contain data on altitude, monetary poverty, or natural region. This data was added to each deceased according to their district of origin. We perform a list of 1854 districts of our country of these three variables. The data was obtained from the National Institute of Statistics and Informatics of Peru (INEI in Spanish). Using a search and assignment formula in Microsoft Excel spreadsheet; data was transferred to each subject.

#### Analysis plan and sample size power

Fist of all, we evaluated the presence of extreme values; recoded texts to prespecified codes and generated new variables as described above [22].

We calculated the proportion of deaths from COVID-19 according to the altitude of area of origin. Both variables were described according to sociodemographic characteristics and underlying cause of death. We evaluated the difference between proportions according to Pearson’s Chi-square test, considering a significance level of 0.05.

Considering that the data presents a geographical hierarchical pattern and people shared common characteristics according to geography. And it is a potential limitation for the assumption of independence of observations. We carried out a multilevel model considering the territorial level of province and natural region. Likewise, it was verified whether the multilevel model would contribute more than the single level model by verifying whether the random disturbance around the constant is zero. [23] Then, we carried out a Poisson regression analysis with robust variance to estimate the risk ratio of death and its relationship with the altitude of residence.We verify the absence of overdispersion using deviance and degrees of freedom. We built a first model adjusted for age and sex. Then a second model, in which we add comorbidities. And a third model, in which we add the contextual variable monetary poverty to the previous model. Finally, we did a sensitivity analysis to evaluate the change in the proportion of deaths every 100, 250, 500 and 1000 m. We presented all RRs with 95% confidence interval.[24] For data analysis, we used Stata v18 (StataCorp, College Station, Texas, USA).

We performed power evaluation to evaluate differences between the proportion of three independent groups. For this we use the artbin formula of Stata 18 program. For the calculation we consider the proportion of death due to COVID of each altitude category, the sampling ratio of each group considering the first group as 1 and the other two groups based on this. a two-tailed 5% level of significance. Obtaining a power of 97.5%.[25] Formula details can be seen in Supplementary Table 4.

#### Ethics

We use publicly accessible data from the website of the Ministry of Health of Peru. No records contained individually identifying data. The research protocol was reviewed and approved by the Institutional ethics and Research Committee of the Universidad Científica del Sur under code CONSTANCIA N°49-CIEI-CIENTIFICA-2021.

## Results

We selected deaths that occurred between March and December 2020 that comprised 188 478 subjects. After excluding subjects without DM, deaths abroad, incomplete data, or unknown cause of death; we finally included 16 406 deaths. Of which 5623 died from COVID-19 (**Fig 1**).

**Fig 1.**
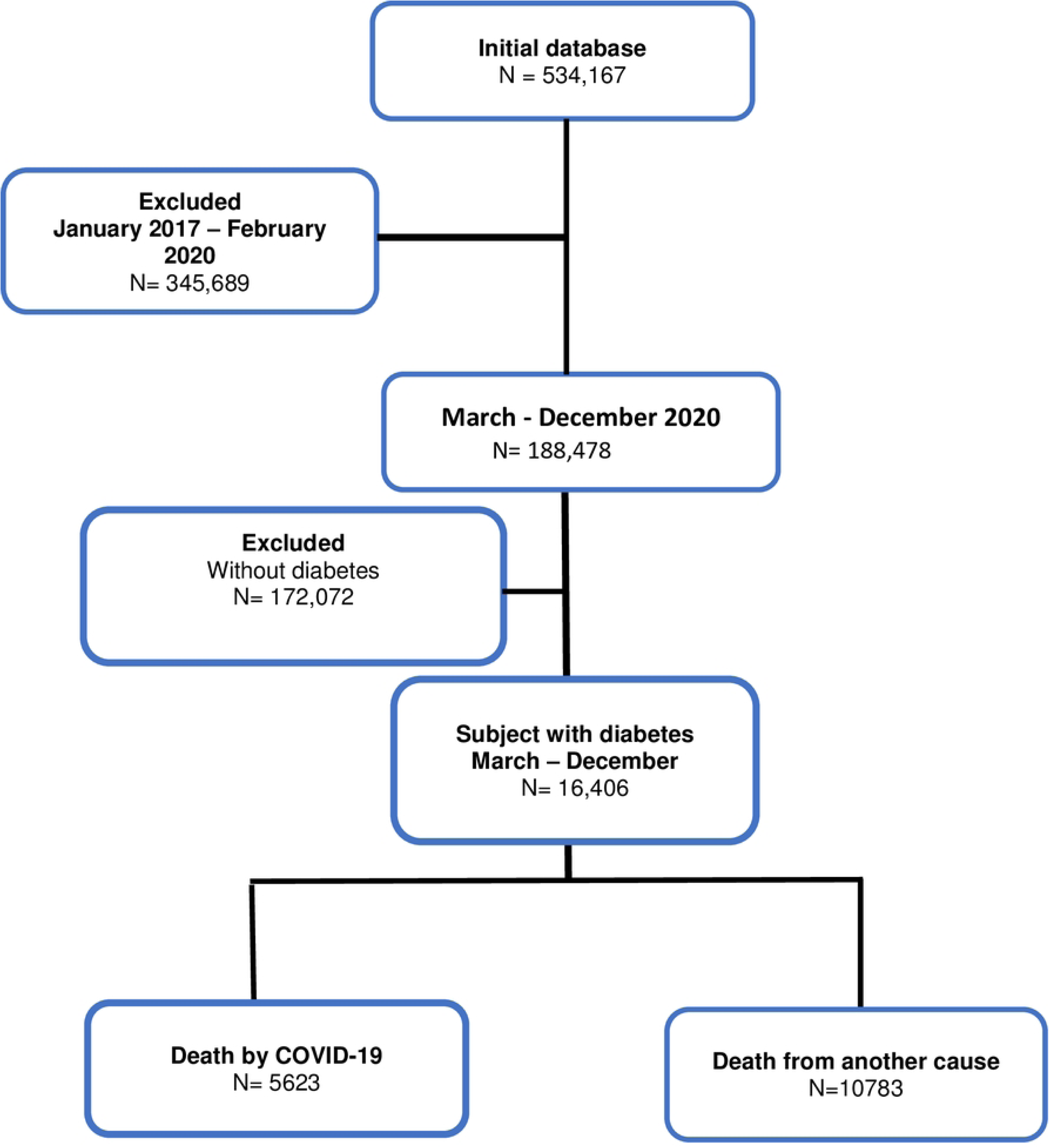
Selection Flow Chart.

### Characteristics of deaths with DM

Of the 16406 deaths with DM, 56% were men and 75% were older than 60 years. Only 6.9% didn’t have insurance, 40% died at home and the region with the highest mortality was Lima (42%). In addition, 75% of deaths come from areas of the quintile with the lowest national percentage of monetary poverty. Likewise, 9.7% came from areas with an altitude above 2 500 m Approximately, 52.3% presented an infectious disease as the basic cause of death and 43.1% a non-communicable cause. In relation to comorbidities, 29.6% reported high blood pressure, 3% obesity and 3.8% cancer of any kind. 40.4% subjects were positive for SARS CoV-2, which included 25.3% as confirmed diagnosis and 9.4% as probable diagnosis. (Table 1)

**Table 1.** Demographic characteristics of deceased with diabetes mellitus according to altitude of origin in Peruvians March - December 2020.

|  | Total | (%) | <1500 m | (%) | 1500 - 2499 m | (%) | ≥ 2500 m | (%) | Valor p |
| --- | --- | --- | --- | --- | --- | --- | --- | --- | --- |
| **General** | 16406 |  | 13649 |  | 1159 |  | 1598 |  |  |
| **Sex** |  |  |  |  |  |  |  |  | <0.001 |
| Female | 7225 | (44.0) | 5917 | (43.3) | 524 | (45.2) | 784 | (49.1) |  |
| Male | 9181 | (55.9) | 7732 | (56.6) | 635 | (54.8) | 814 | (50.9) |  |
| **Age group** |  |  |  |  |  |  |  |  |  |
| Media ± SD | 68.2 ± 14.1 |  | 68.3 ± 14.1 |  | 69.4 ± 14.0 |  | 64.6 ± 12.6 |  |  |
| 0 – 11.9 | 18 | (0.1) | 17 | (0.1) | 0 | (0) | 1 | (0.1) |  |
| 12 – 17.9 | 13 | (0.1) | 12 | (0.1) | 1 | (0.1) | 0 | (0) |  |
| 18 – 29.9 | 133 | (0.8) | 104 | (0.7) | 11 | (0.9) | 18 | (1.1) |  |
| 30 – 59.9 | 3924 | (23.9) | 3216 | (23.6) | 259 | (22.4) | 449 | (28.1) |  |
| ≥ 60 years | 12317 | (75.1) | 10299 | (75.5) | 888 | (76.6) | 1130 | (70.7) |  |
| **Education level** |  |  |  |  |  |  |  |  |  |
| Illiterate | 1424 | (8.7) | 1049 | (7.7) | 109 | (9.4) | 266 | (16.7) | <0.001 |
| Primary | 4141 | (25.2) | 3366 | (24.7) | 310 | (26.7) | 465 | (29.1) |  |
| Secondary | 4280 | (26.1) | 3612 | (26.5) | 293 | (25.3) | 375 | (23.5) |  |
| Higher | 2025 | (12.3) | 1601 | (11.7) | 182 | (15.7) | 242 | (15.1) |  |
| No registration | 4536 | (27.7) | 4021 | (29.4) | 265 | (22.9) | 250 | (15.6) |  |
| **Health insurance** |  |  |  |  |  |  |  |  |  |
| General population | 7543 | (46.0) | 6109 | (44.8) | 451 | (38.9) | 983 | (61.5) | <0.001 |
| Employee | 6542 | (39.9) | 5577 | (40.9) | 525 | (45.3) | 440 | (27.5) |  |
| Armed Forces and Police | 491 | (3.00) | 428 | (3.1) | 27 | (2.3) | 36 | (2.3) |  |
| Private / others | 690 | (4.2) | 595 | (4.4) | 35 | (3.0) | 60 | (3.7) |  |
| Uninsured | 1140 | (6.9) | 940 | (6.9) | 121 | (10.4) | 79 | (4.9) |  |
| **Place of death** |  |  |  |  |  |  |  |  |  |
| Healthcare center | 9861 | (60.1) | 8213 | (60.2) | 705 | (60.8) | 943 | (59.0) | <0.001 |
| Home | 6233 | (38.0) | 5223 | (38.3) | 397 | (34.3) | 613 | (38.3) |  |
| Public road/in transit | 300 | (1.8) | 201 | (1.5) | 57 | (4.9) | 42 | (2.6) |  |
| Unknown | 12 | (0.1) | 12 | (0.1) | 0 | (0.0) | 0 | (0.0) |  |
| **Natural region** |  |  |  |  |  |  |  |  |  |
| Lima-Callao | 6872 | (41.9) | 6872 | (50.4) | 0 | (0.0) | 0 | (0.0) | <0.001 |
| Rest of coast | 4968 | (30.3) | 4968 | (36.4) | 0 | (0.0) | 0 | (0.0) |  |
| Sierra | 3001 | (18.3) | 381 | (2.8) | 1022 | (88.2) | 1598 | (100.0) |  |
| Jungle | 1565 | (9.5) | 1428 | (10.4) | 137 | (11.8) | 0 | (0.0) |  |
| **Poverty quintile <sup>a</sup>** |  |  |  |  |  |  |  |  |  |
| Quintile 5 (Greater poverty) | 197 | (1.2) | 45 | (0.3) | 37 | (3.2) | 115 | (7.2) | <0.001 |
| Quintile 4 | 347 | (2.1) | 102 | (0.8) | 43 | (3.7) | 202 | (12.6) |  |
| Quintile 3 | 757 | (4.6) | 557 | (4.1) | 12 | (1.0) | 188 | (11.7) |  |
| Quintile 2 | 2877 | (17.5) | 2332 | (17.1) | 66 | (5.7) | 479 | (30.0) |  |
| Quintile 1 (Less poverty) | 12228 | (74.5) | 10613 | (77.8) | 1001 | (86.4) | 614 | (38.4) |  |
| **Basic cause of death** |  |  |  |  |  |  |  |  |  |
| Infectious | 8573 | (52.3) | 7176 | (52.6) | 613 | (52.9) | 784 | (49.1) | <0.001 |
| Not communicable | 7075 | (43.1) | 5851 | (42.9) | 456 | (39.3) | 768 | (48.1) |  |
| External | 487 | (2.9) | 392 | (2.9) | 59 | (5.1) | 36 | (2.3) |  |
| Not determined | 271 | (1.7) | 230 | (1.7) | 31 | (2.7) | 10 | (0.6) |  |
| **SARS-CoV-2** |  |  |  |  |  |  |  |  |  |
| Negative | 9770 | (59.6) | 8029 | (58.8) | 706 | (60.9) | 1035 | (64.8) | <0.001 |
| Positive <sup>b</sup> | 6636 | (40.4) | 5620 | (41.2) | 453 | (39.1) | 563 | (35.2) |  |
| Confirmed | 4159 | (25.3) | 3455 | (25.3) | 271 | (23.4) | 433 | (27.1) |  |
| Probable | 1548 | 9.4 | 1334 | 9.8 | 139 | 12.0 | 75 | 4.7 |  |
| Other code | 929 | 5.7 | 831 | 6.1 | 43 | 3.7 | 45 | 3.4 |  |
| **Comorbidity** |  |  |  |  |  |  |  |  |  |
| **Hypertension** |  |  |  |  |  |  |  |  |  |
| Yes | 4849 | (29.6) | 4326 | (31.7) | 249 | (21.5) | 274 | (17.2) | <0.001 |
| No | 11557 | (70.4) | 9323 | (68.3) | 910 | (78.5) | 1324 | (82.8) |  |
| **Obesity** |  |  |  |  |  |  |  |  |  |
| Yes | 490 | (3.0) | 405 | (3.0) | 40 | (3.4) | 45 | (2.8) | 0.594 |
| No | 15916 | (97.0) | 13244 | (97.0) | 1119 | (96.6) | 1553 | (97.2) |  |
| **Cancer** |  |  |  |  |  |  |  |  |  |
| Yes | 627 | (3.8) | 534 | (3.9) | 42 | (3.6) | 51 | (3.2) | 0.340 |
| No | 15779 | (96.2) | 13115 | (96.1) | 1117 | (96.4) | 1547 | (96.8) |  |
| **Coronary Heart Disease** |  |  |  |  |  |  |  |  |  |
| Yes | 2735 | (16.7) | 2448 | (17.9) | 182 | (15.7) | 105 | (6.6) | <0.001 |
| No | 13671 | (83.3) | 11201 | (82.1) | 977 | (84.3) | 1493 | (93.4) |  |
| **Stroke** |  |  |  |  |  |  |  |  |  |
| Yes | 799 | (4.9) | 688 | (5.0) | 48 | (4.1) | 63 | (3.9) | 0.076 |
| No | 15607 | (95.1) | 12961 | (95.0) | 1111 | (95.9) | 1535 | (96.1) |  |

Source: National System of Deaths from March to December 2019 and 2020.
<sup>a</sup> People residing in households whose per capita spending is insufficient to acquire a basic basket of food and non-food (INEI)
<sup>b</sup> COVID positive: ICD10 (U07.1, U07.2, B34.X or J97.X) or by written diagnosis (“COVID”, “Coronavirus”, “SARS CoV-2”). COVID Negative: Absence of any of the words in the previous definition.

### Characteristics of deaths with DM above 2500 m

In subjects living above 2 500 m a similar proportion of men and women was observed, unlike low-lying areas, in which men predominated (p<0.001). A slightly higher proportion of middle-aged subjects and lower proportion of older adults was observed (p<0.001). These patients had a higher proportion of illiterates and came from areas with the highest monetary poverty quintile. Non-communicable causes of death predominated more frequently. Regarding comorbidities, lower arterial hypertension and lower coronary disease were observed, but obesity and cancer of any type were similar. The place of death at home or in a health center was like that in areas of lower altitude (Table 1).

### Proportion of death from COVID-19

The proportion of deaths from COVID-19 was 34.3% (95% CI 33.6–35.1). According to altitude, it was 34.8% (95% CI; 34.0 – 35.6) below 1 500 m. 34.6% (95% CI; 31.8 – 37.4) between 1 500-2 499 m and 29.6% (95% CI; 27.4 – 31.9) above 2 500 m.

The proportion of deaths from COVID-19 was higher in males than in females (37.7%; vs 29.9%; p<0.001). The highest proportion occurred in the middle age compared to other age groups (40.5% between 30 to 59 years old, versus 32.4% in those older than 60 years; p<0.001). A higher proportion was also observed with a higher level of instruction (15.2% in illiterate or 16.9% in primary school vs. 28.2% in secondary school and 27.2% in higher education; p<0.001).

Regarding health insurance, 48.1% with military or police insurance died from COVID-19, 35.7% with General Population Health Insurance and 33.9% with Employee Health insurance (p<0.001). Regarding the place of death, 53.2% who died in health centers was due to COVID-19 and 5.1% at home (p<0.001).

The coastal area had the highest percentage of deaths from COVID 19, much more so in the area outside of Lima/Callao. 37.1 vs 33.8. In descending order, there were the mountains and the jungle. (p<0.001).

The frecuency of death by poverty quintile was U-shaped. In quintile 1 (less poverty), 35.7% died from COVID-19, falling to 31.5% in quintile 2 and then rising progressively in quintile 5 with 36.6%. (Table 2)

**Table 2.**
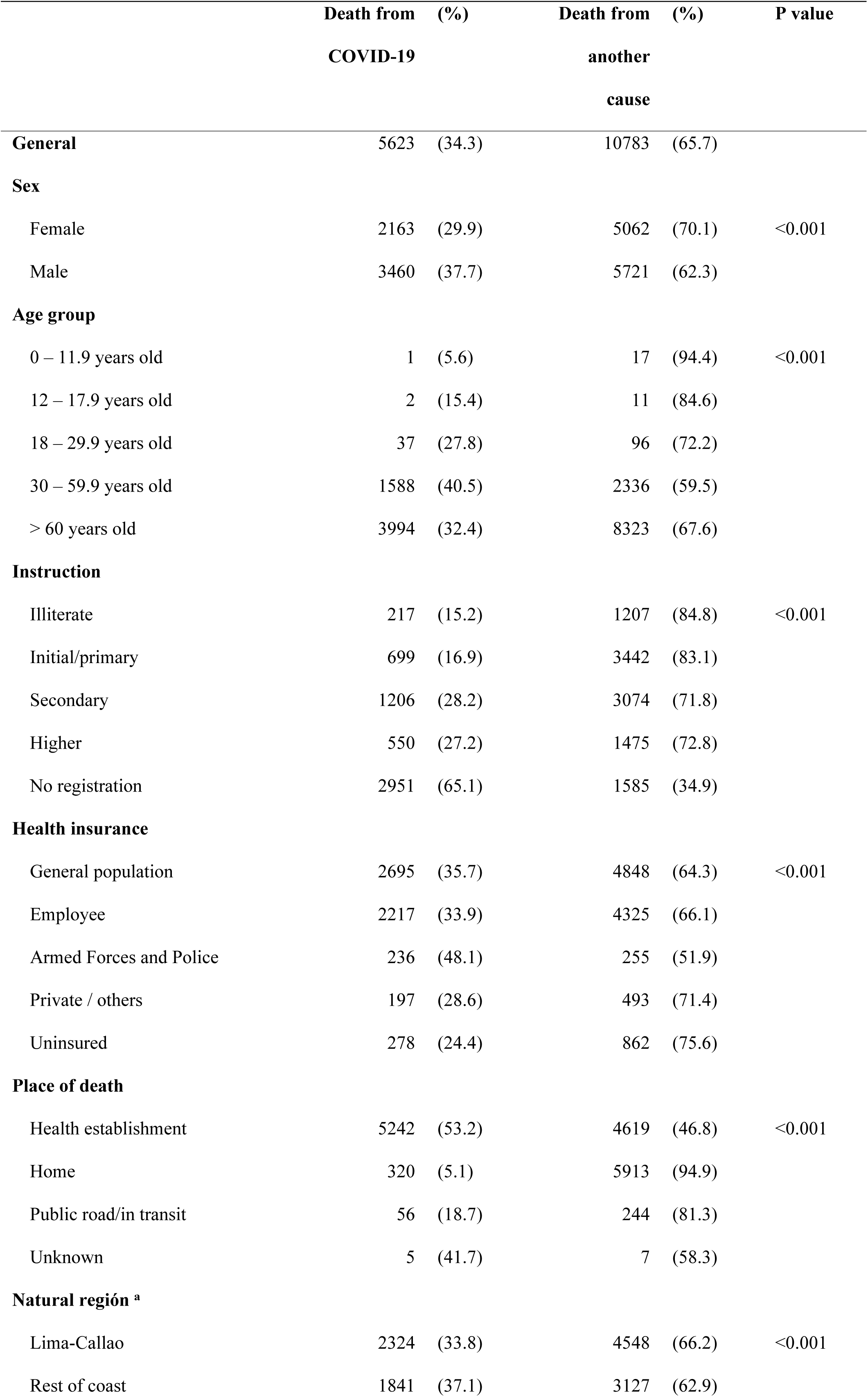
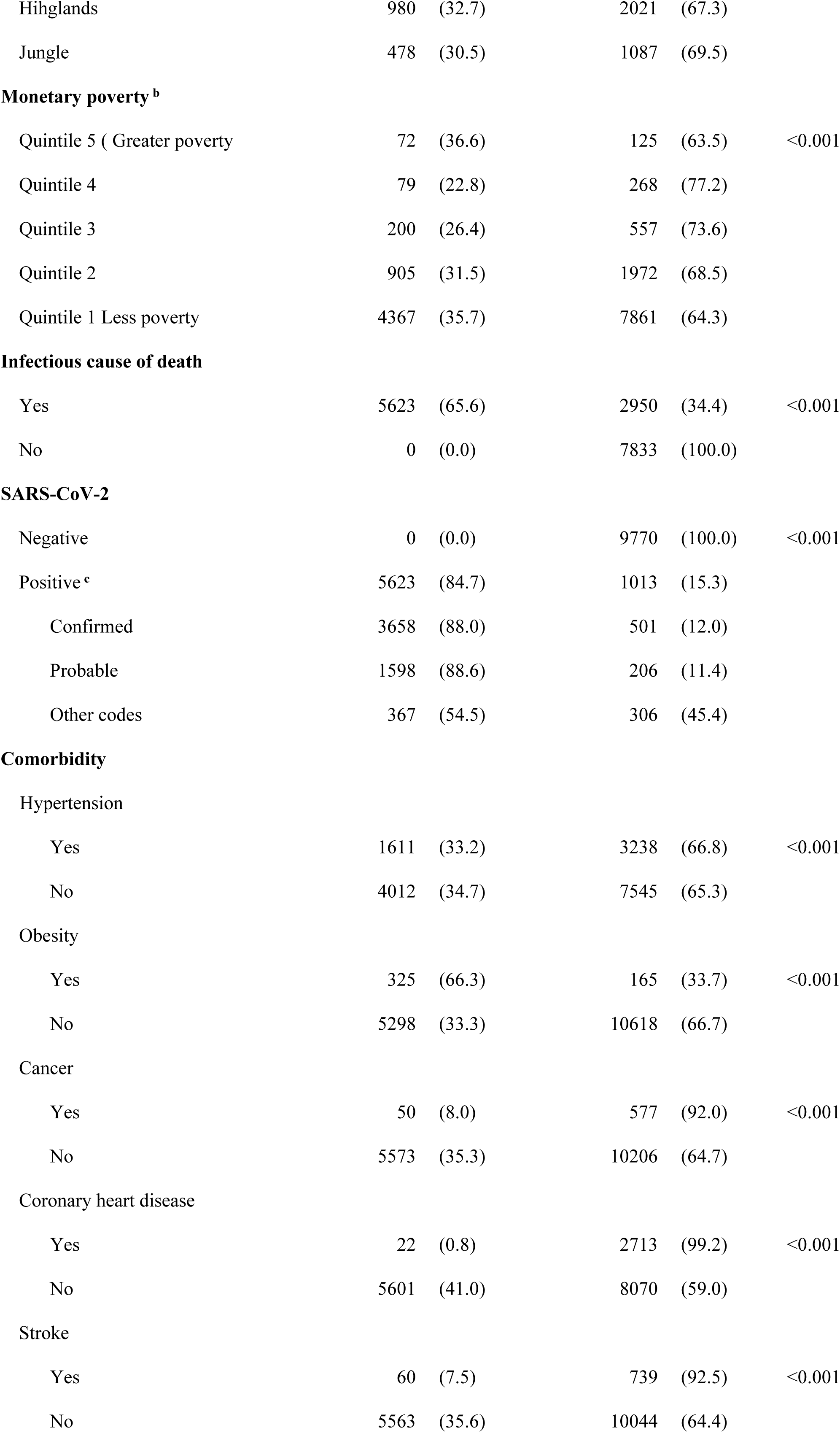
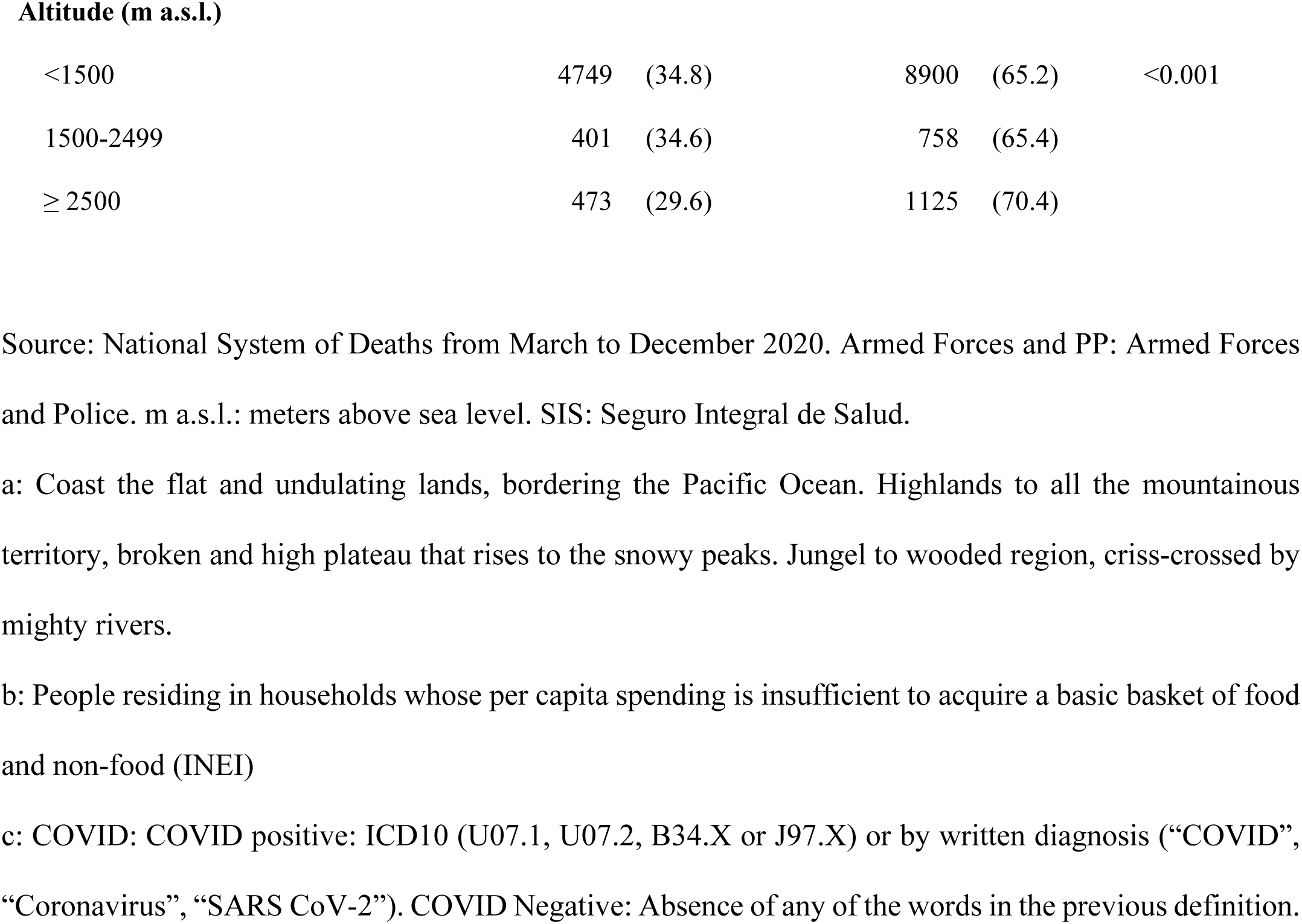
Proprotion of death from COVID-19 as the underlying causeaccording to demographic characteristics in deceased with diabetes mellitus.

### Risk ratios of death from COVID-19 according to altitude

There were 20% fewer deaths of those from an area above 2 500 m compared to lower altitude areas adjusted for age, sex, high blood pressure, obesity, cancer, coronary heart disease, stroke and monetary poverty (RR 0.80; 95% CI 0.75-0.88; p <0.001). (**Table 3** and Supplementary table 5)

**Table 3.** Crude and adjusted risk ratio of death from COVID-19 as the underlying cause according to altitude in deceased Peruvians with diabetes mellitus.

|  | Crude analysis |  | Ajusted model A |  | Ajusted model B |  | Ajusted model C |  |
| --- | --- | --- | --- | --- | --- | --- | --- | --- |
|  | RR (95% CI) | p-value | RR (95% CI) | p-value | RR (95% CI) | p-value | RR (95% CI) | p-value |
| **Altitude (m a.s.l.)** |  |  |  |  |  |  |  |  |
| <1500 | 1.00 |  | 1.00 |  | 1.00 |  | 1.00 |  |
| 1500- 2499 | 0.87 (0.71 – 1.09) | 0.242 | 0.90 (0.73 – 1.11) | 0.341 | 0.90 (0.74 . 1.11) | 0.351 | 0.90 (0.75 – 1.09) | 0.290 |
| ≥ 2500 | 0.79 (0.69 – 0.91) | <0.001 | 0.79 (0.69 – 0.91) | <0.001 | 0.76 (0.67 – 0.86) | <0.001 | 0.80 (0.70 – 0.91) | <0.001 |

RR: Risk Ratio
Model A: Adjusted for age and sex. Model B: Adjusted for age, sex, high blood pressure, obesity, cancer, coronary heart disease, stroke. Model C: Adjusted for age, sex, high blood pressure, obesity, cancer, coronary heart disease, stroke and monetary poberty

### Sensitivity analysis

We performed a sensitivity analysis every 100, 250, 500 and 1 000 m and observed a constant decrease as altitude increases. The proportion of deaths from COVID-19 decreased by 0.8% every 100 m a.s.l. (RR 0.992; 95% CI 0.988-0.996, p=<0.001); 1.8% every 250 m a.s.l. (RR 0.982; 95% CI 0.972-0.992; p<0.001); 4.0% every 500 m a.s.l (RR 0.960; 95% CI 0.940-0.981; p<0.001) or 7.1% every 1000 m a.s.l. (RR 0.929; 95% CI 0.893-0.967; p<0.001). All RR also were adjusted for age, sex, high blood pressure, obesity, cancer, coronary heart disease, stroke and monetary poverty (**Table 4** and Supplementary tables 6,7,8 and 9).

**Table 4.** Sensitivity analysis of altitude in deceased Peruvians with diabetes mellitus.

|  | Crude analysis |  | Adjusted model A |  | Adjusted model B |  | Adjusted model C |  |
| --- | --- | --- | --- | --- | --- | --- | --- | --- |
|  | RR (95% CI) | p-value | RR (95% CI) | p-value | RR (95% CI) | p-value | RR (95% CI) | p-value |
| **Altitude (m** |  |  |  |  |  |  |  |  |
| **a.s.l)** |  |  |  |  |  |  |  |  |
| Δ 100 m | 0.992 (0.988 – 0.997) | 0.002 | 0.993 (0.988 – 0.997) | 0.002 | 0.991 (0.987 – 0.995) | <0.001 | 0.992 (0.988 -. 0.996) | <0.001 |
| Δ 250 m | 0.982 (0.971 – 0.993) | 0.002 | 0.983 (0.972 – 0.994) | 0.002 | 0.978 (0.968 – 0.988) | <0.001 | 0.982 (0.972 -. 0.992) | <0.001 |
| Δ 500 m | 0.960 (0.939 – 0.982) | <0.001 | 0.962 (0.940 – 0.983) | 0.002 | 0.953 (0.933 – 0.973) | <0.001 | 0.960 (0.940 - 0.981) | <0.001 |
| Δ 1000 m | 0.9310 (0.890 – 0.973) | 0.002 | 0.933 (0.892 – 0.975) | 0.002 | 0.916 (0.880 – 0.954) | <0.001 | 0.929 (0.893 - 0.967) | <0.001 |

Model A: Adjusted for age and sex. Model B: Adjusted for age, sex, high blood pressure, obesity, cancer, coronary heart disease, stroke. Model C: Adjusted for age, sex, high blood pressure, obesity, cancer, coronary heart disease, stroke and monetary poberty.

## Discussion

Altitude modifies glucose metabolic in patients with DM improving insulin sensitivity and potentially protecting against complications. Our study found that in those deceased with DM, one out of three died due to COVID-19 and one out of 10 deaths occurred above 2 500 m. Likewise, the proportion of deaths from COVID-19 above 2500 m was 20% lower compared to lower altitudes.

Many of the countries in the world were affected at the first level of care by COVID-19 during the first wave. Facility closures, staff shortages or supply problems affected one out of four emergency services in up to 25% of countries [26]. Emergency care, preventive-promotional care, and outpatient consultations were limited leading to the strengthening of telehealth and telemedicine. [27]

In Peru, a quarter of the Peruvian population lives above 2 500 m [28] and there is a greater population located in the capital cities where overcrowding and number of vehicles and greater pollution that facilitated SARS-CoV-2 expansion [29] The return of the immobilized population in the big cities to their places of origin raised the rates of the virus in the high Andean areas. However, this lower trend in rates was maintained when adjusted for population. Similar to our study, a report in Mexico compared deceased with diabetes according altitude of residence. Those who lived above 1 500 presented fewer cases of COVID pneumonia and a lower proportion of deaths. However, they were at higher risk than the population without diabetes.[30]. Other clinical series based on general population maintain this trend of lower proportion of death at higher altitudes, adjusting for population density and poverty level. [31]

It is postulated that under conditions of hypoxia, the production of hypoxia-inducible transcription factor 1 (HIF1) is increased, so that ACE increases and stimulates the expression of angiotensin II (ACE2), which regulates AT1 receptors, generating a lower expression of ACE2. Fewer ACE2 receptors would mean a lower viral load and decrease of SARS-CoV-2 infection.[32] Likewise, the increase expression of erythropoietin presents a cytoprotective action, reducing inflammation and improving the microvascular injury of cytotoxins in the blood [33]

On another hand, some reports have demonstrated the absence of this clinical impact. [34]. However, in patients with diabetes mellitus, this is not clear. Chronic exposure to high altitude results in low glucose levels mediated by improved insulin sensitivity and increased peripheral glucose uptake. The reasons for these adaptations are still unknown. Likewise, an inverse relationship between altitude with diabetes and obesity is well documented. Being the result of genetic and physiological adaptations, due to hypoxia in the glucose metabolism. important factors are also diet and lifestyle patterns.[12]

Analyzing this relationship by subgroups. In men, an increased risk of death from COVID-19 could be due to a greater number of ACE receptors, however lower rate was observed at higher altitudes. In women, immunosuppressive effect of estrogens also led to lower mortality [35]. According to age, case fatality rate have higher numbers in older age groups. However, the percentage of deaths from COVID-19 in each age group was higher in the middle age group because this age group presented an exposure without having adequate security measures to carry the daily monetary income due to prolonged closure . [36]

### Importance in public health

Peru is one of the first countries with the highest mortality from SARS-CoV-2 in Latin America and the world. Non-communicable diseases were the most prevalent in deceased diabetics. It is important to recognize the limitations that arose during the first wave and its impact.

Diabetes control was affected by the closure of primary care centers [37]. One study reported that glycemic control and high fasting blood sugar worsened during the SARS-CoV-2 outbreak at the big cities which caused a higher incidence of complications and poor control of the disease [38]. Likewise, this study contribute to expanding knowledge of the effects of altitude with respect to mortality in people with diabetes mellitus in a context of a highly contagious and virulent infectious disease.

Also, we recommended a permanent awareness and training campaign is suggested for all medical personnel to correctly fill out the death certificate, due to the deficiencies found now and in previous studies that limit their correct interpretation [39].

### Limitations and strengths

One of our limitations was the poor registration of the death certificates of the basic cause of death and the use of the ICD-10. Due to serological tests, there was a high proportion of false negatives and were the main diagnostic method during the first wave in Peru. The diabetes mellitus assignment may have been underrepresented, because there are only six fields for that. The first four for the causes and two for the comorbidities. The altitude was obtained according to the district of origin of the death based on the official government records however may not be precise in those who migrated or have a stable residence at another altitude. The design does not allow a causal relationship to be established with high certainty between the variables, but it does allow the influence of the basic sociodemographic variables to be analyzed. Another limitation was not being able to include clinical or therapeutic variables that potentially influence the association. As well as being able to differentiate those cases of hyperglycemia due to stress caused by SARS-CoV-2 infection, which may overestimate some results.

On the other hand, one of our strengths is that the SINADEF has registered all the deceased at the national level, obtaining a better analysis of region, province and district level. We perform a multilevel analysis based in province and natural region. As well as a sensitivity analysis using different altitude increments.

### Conclusion

During the first wave, the proportion of deaths from COVID-19 of people residing above 2 500 m was 20% lower than in areas of lower altitude, adjusted for individual and socioeconomic factors. Its influence is also shown as the altitude changes every 100, 250, 500 and 1000 m a.s.l., through multilevel analysis. In high Andean areas, there is greater insulin sensitivity which may be a protective factor against complications. The study contribute to expanding knowledge of the effects of altitude with respect to mortality in people with diabetes mellitus in a context of a highly contagious and virulent infectious disease.

## Data Availability

All relevant data are within the manuscript and its Supporting Information files.

## Supporting information

**S1 table. Codes and words used to search for the diagnosis in the database.**

**S2 table. Codes or texts used within the four groups**

**S3 table. Lower and upper limit of the monetary poverty index of each quintile of the sample.**

**S4 table. Calculation of sample power**

**S5 table. Full regression analysis of death from COVID-19 as the underlying cause according to altitude in deceased Peruvians with diabetes mellitus.**

**S6 table. Full regression analysis of death from COVID-19 as the underlying cause according to altitude in deceased Peruvians with diabetes mellitus each 100 m a.s.l**

**S7 table. Full regression analysis of death from COVID-19 as the underlying cause according to altitude in deceased Peruvians with diabetes mellitus each 250 m a.s.l**

**S8 table. Full regression analysis of death from COVID-19 as the underlying cause according to altitude in deceased Peruvians with diabetes mellitus each 500 m a.s.l**

**S9 table. Full regression analysis of death from COVID-19 as the underlying cause according to altitude in deceased Peruvians with diabetes mellitus each 1000 m a.s.l**

## Notes

### Competing Interest Statement

The authors have declared no competing interest.

### Funding Statement

The author(s) received no specific funding for this work.

### Author Declarations

We use publicly accessible data from the website of the Ministry of Health of Peru. No records contained individually identifying data. The research protocol was reviewed and approved by the Institutional ethics and Research Committee of the Universidad Científica del Sur under code N°49-CIEI-CIENTIFICA-2021.

